# Nonpharmaceutical interventions for pandemic COVID-19: A cross-sectional investigation of US general public beliefs, attitudes, and actions

**DOI:** 10.1101/2020.04.26.20078618

**Authors:** Bella Nichole Kantor, Jonathan Kantor

## Abstract

Nonpharmaceutical interventions (NPIs) represent the primary mitigation strategy for pandemic COVID-19. Despite this, many government agencies and members of the general public may be resistant to NPI adoption. We sought to understand public attitudes and beliefs regarding various NPIs and self-reported adoption of NPIs, and explore associations between NPI performance and the baseline characteristics of respondents. We performed a cross-sectional age-, sex-, and race- stratified survey of the general US population. Of the 1,005 respondents, 37% (95% CI 34.0, 39.9) felt that NPIs were inconvenient, while only 0.9% (95% CI 0.3, 1.5) of respondents believed that NPIs would *not* reduce their personal risk of illness. Respondents were most uncertain regarding the efficacy of mask and eye protection use, with 30.6% and 22.1%, respectively, unsure whether their use would slow disease spread. On univariate logistic regression analyses, NPI adherence was associated with a belief that NPIs would reduce personal risk of developing COVID-19 (OR 3.06, 95% CI [1.25, 7.48], p=0.014) and with a belief that the NPIs were *not* difficult to perform (OR 1.79, 95% CI [1.38, 2.31], p<0.0001). Respondents were compliant with straightforward, familiar, and heavily-encouraged NPI recommendations such as hand-washing; more onerous approaches, such as avoiding face touching, disinfecting surfaces, and wearing masks or goggles, were performed less frequently. NPI non-adherence is associated with both outcome expectations (belief that NPIs are effective) and process expectations (belief that NPIs are not overly inconvenient); these findings have important implications for designing public health outreach efforts, where the feasibility, as well as the effectiveness, of NPIs should be stressed.

## Introduction

Nonpharmaceutical interventions (NPIs) have emerged as a first line of protection and mitigation in the face of pandemic SARS-CoV-2 infection, particularly given evidence suggesting the efficacy of such interventions in previous pandemics.^1,2^ Since modern NPIs were adopted over a century ago during the 1918–1919 flu pandemic, much of the public debate has remained unchanged, centering on the efficacy and burdensomeness of NPIs, and their potential for broader effects on morale and economic stability.^3,4^

Few studies have evaluated public perceptions of NPIs in the context of influenza pandemics,^5,6^ and none have addressed this issue in the context of the present COVID-19 pandemic. Pandemic responsiveness is contingent on individuals eschewing their normal daily behaviors; thus a small number of refusers may drive—and social media may further exacerbate—such behaviors. Some have suggested that NPI adherence is improved with improved communication; that is, NPI non-adherence is a function of a knowledge gap.^7^ Yet data from behavioral research suggests that non-compliance with expert recommendations is generally *not* a function of a lack of knowledge per se.^8–10^

Understanding whether outcome expectations (a perception of *efficacy*) affects NPI adherence is critical; if there is a knowledge gap in appreciating that NPIs are effective, it could be addressed through outreach efforts. Conversely, if NPI non-adherence is a function of process expectations (concerns that performing NPIs is too *onerous*), then outreach efforts could be focused on mitigating these perceptions rather than highlighting the potential to reduce disease spread.

We therefore sought to understand public attitudes and beliefs regarding various NPIs and self-reported adoption of NPIs, and explore associations between NPI performance and the baseline characteristics of respondents. These data may help inform public health efforts, as better understanding the drivers of refusal to engage in NPIs will help tailor messaging appropriately and ideally increase the chances of encouraging behavioral changes that may ultimately result in reduced disease transmission.

## Methods

We developed a cross-sectional online survey of the general US population after iterative pilot testing. This study was deemed exempt by the Ascension Health institutional review board. The survey was prepared on the Qualtrics platform (Qualtrics Corp, Provo, Utah) and distributed to a representative US sample stratified by age, sex, and race, through Prolific Academic (Oxford, United Kingdom), a platform for academic survey research.^11^ Respondents were rewarded with a small payment (<US$1). Subjects provided consent and were allowed to terminate the survey at any time, and all responses were confidential.

Baseline responses to survey questions were recorded (Supplemental file), and demographic information was self-reported by respondents. Responses to a range of questions regarding attitudes to the COVID-19 pandemic, fears, worry, and NPI beliefs and actions were collected using Likert scales.

T-tests and chi-squared tests were used as appropriate for baseline continuous and categorical variables. Subgroup comparisons of non-normally distributed data were performed using the Kruskal Wallis test. Univariate logistic regression odds ratios of association were assessed between the dependent variable of NPI adherence, defined as those who engaged, on average, in each NPI always or most of the time, and baseline characteristics and attitudes. Statistical analyses were performed using Stata 13 for Mac (College Station, Texas).

## Results

Of the 1,020 subjects who were recruited, 1,005 finished the survey, yielding a completion rate of 98.5%. The mean (SD) age of respondents was 45 (16), and 494 (48.8%) of the respondents were male; baseline respondent characteristics are outlined in Table 1.

**Table 1.** Demographic and baseline characteristics of respondents, overall and by social distancing adherence and shelter in place status.

|  |  | *Social distancing* |  | *Under a shelter in place order* |  |
| --- | --- | --- | --- | --- | --- |
| **Characteristic** | **Total** | **Always** | **Not Always** | **Yes** | **No** |
| **Overall** | 1005 (100) | 736 (72.2) | 284 (27.8) | 681 (66.8) | 389 (33.2) |
| **Sex<sup>†</sup>** |  |  |  |  |  |
| **Men** | 494 (48.8) | 347 (47.2) | 147 (53.3) | 310 (46.1) | 184 (54.3) |
| **Women** | 518 (51.2) | 389 (52.9) | 129 (46.7) | 363 (53.9) | 155 (45.7) |
| **Age, y** |  |  |  |  |  |
| **18-30** | 250 (24.5) | 171 (23.2) | 79 (27.8) | 165 (24.2) | 85 (25.1) |
| **31-40** | 204 (20.0) | 152 (20.7) | 52 (18.3) | 139 (20.4) | 65 (19.2) |
| **41-50** | 146 (14.3) | 108 (14.7) | 38 (13.4) | 100 (14.7) | 46 (13.6) |
| **51-60** | 198 (19.8) | 151 (20.5) | 47 (16.6) | 130 (19.1) | 68 (20.1) |
| **>60** | 222 (21.8) | 154 (20.9) | 68 (23.9) | 147 (21.6) | 75 (22.1) |
| **Education level<sup>\*</sup>** |  |  |  |  |  |
| **< High school** | 11 (1.1) | 4 (0.5) | 7 (2.6) | 8 (1.2) | 3 (0.9) |
| **High school** | 117 (11.7) | 77 (10.5) | 40 (15.0) | 67 (10.1) | 50 (14.8) |
| **Some college** | 228 (22.8) | 166 (22.6) | 62 (23.2) | 149 (22.4) | 79 (23.4) |
| **Associates** | 103 (10.3) | 72 (9.8) | 31 (11.6) | 66 (9.9) | 37 (11.0) |
| **Bachelor's** | 358 (35.7) | 272 (37.0) | 86 (32.2) | 246 (37.0) | 112 (33.2) |
| **Graduate** | 185 (18.5) | 144 (19.6) | 41 (15.4) | 129 (19.4) | 56 (16.6) |
| **Employment status** |  |  |  |  |  |
| **Full time** | 461 (45.2) | 339 (46.1) | 122 (43.0) | 303 (44.5) | 158 (46.6) |
| **Part time** | 170 (16.7) | 118 (16.0) | 52 (18.3) | 115 (16.9) | 55 (16.2) |
| **Not employed** | 389 (38.1) | 279 (37.9) | 110 (38.7) | 263 (38.6) | 127 (37.2) |
| **Religious** |  |  |  |  |  |
| **Yes** | 387 (37.9) | 279 (37.9) | 108 (38.0) | 252 (37.0) | 135 (39.8) |
| **No** | 543 (53.2) | 391 (53.1) | 152 (53.5) | 361 (53.0) | 182 (53.7) |
| **Ambivalent** | 90 (8.8) | 66 (9.0) | 24 (8.5) | 68 (10.0) | 22 (6.5) |
| **Income** |  |  |  |  |  |
| **<\$10,000** | 167 (16.4) | 121 (16.4) | 46 (16.2) | 115 (16.9) | 52 (15.3) |
| **\$10,000-\$30,000** | 234 (22.9) | 169 (23.0) | 65 (22.9) | 154 (22.6) | 80 (23.6) |
| **\$30,001-\$50,000** | 220 (21.6) | 155 (21.1) | 65 (22.9) | 137 (20.1) | 83 (24.5) |
| **\$50,001-\$80,000** | 201 (19.7) | 151 (20.5) | 50 (17.6) | 131 (19.2) | 70 (20.7) |
| **\$80,001-\$100,000** | 63 (6.2) | 50 (6.8) | 13 (4.6) | 42 (6.2) | 21 (6.2) |
| **\$100,001-\$150,000** | 91 (8.9) | 56 (7.6) | 35 (12.3) | 71 (10.4) | 20 (5.9) |
| **>\$150,000** | 44 (4.3) | 34 (4.6) | 10 (3.5) | 31 (4.6) | 13 (3.8) |
| **Location<sup>†</sup>** |  |  |  |  |  |
| **Urban** | 743 (72.8) | 543 (73.8) | 200 (70.4) | 517 (75.9) | 226 (66.7) |
| **Rural** | 277 (27.2) | 193 (26.2) | 84 (29.6) | 164 (24.1) | 113 (33.3) |

All values are listed as number (%)
<sup>\*</sup> p<0.05 by chi squared test (social distancing)
<sup>†</sup> p<0.05 by chi squared test (shelter in place)

More than 90% of subjects reported using several common NPIs all or most of the time (Table 2). Respondents were most uncertain regarding the efficacy of mask and eye protection use, with 30.6% and 22.1%, respectively, unsure whether their use would slow disease spread. Overall, 37% (34.0, 39.9) of respondents felt that NPIs in general were inconvenient, while only 0.9% (0.3, 1.5) of respondents believed that NPIs in general would *not* reduce their personal risk of illness.

**Table 2.** Nonpharmaceutical intervention performance frequency and belief level.

| NPI | *Performed in Last Week, frequency, n (%)* |  |  |  |  | *Slows the Spread of COVID-19, level of agreement, n (%)* |  |  |  |  |
| --- | --- | --- | --- | --- | --- | --- | --- | --- | --- | --- |
|  | Always | Most of the time | Sometimes | Rarely | Never | Completely agree | Agree | Unsure | Disagree | Disagree completely |
| Hand washing | 776 (77.2) | 188 (18.7) | 29 (2.9) | 9 (0.9) | 3 (0.3) | 871 (86.7) | 124 (12.3) | 9 (0.9) | 0 (0) | 1 (0.1) |
| Hand sanitizer | 355 (35.6) | 192 (19.3) | 222 (22.3) | 95 (9.5) | 132 (13.3) | 722 (71.9) | 224 (22.3) | 45 (4.5) | 7 (0.7) | 6 (0.6) |
| Avoiding handshakes | 875 (87.2) | 67 (6.7) | 42 (4.2) | 9 (0.9) | 10 (1.0) | 819 (81.9) | 164 (16.4) | 13 (1.3) | 2 (0.2) | 2 (0.2) |
| Tissue/ elbow sneeze | 749 (74.6) | 170 (16.9) | 50 (5.0) | 19 (1.9) | 16 (1.6) | 793 (78.9) | 189 (18.9) | 20 (2.0) | 2 (0.2) | 1 (0.1) |
| Avoiding face touching | 247 (24.6) | 356 (35.4) | 282 (28.1) | 86 (8.6) | 34 (3.4) | 748 (74.6) | 207 (20.7) | 42 (4.2) | 5 (0.5) | 1 (0.1) |
| Disinfecting surfaces | 347 (34.7) | 293 (29.3) | 242 (24.2) | 67 (6.7) | 52 (5.2) | 745 (74.2) | 223 (22.2) | 28 (2.8) | 5 (0.5) | 3 (0.3) |
| Wearing mask | 71 (7.1) | 40 (4.0) | 95 (9.5) | 109 (10.9) | 687 (68.6) | 420 (41.9) | 234 (23.4) | 221 (22.1) | 89 (8.9) | 38 (3.8) |
| Wearing eye protection | 77 (7.7) | 45 (4.5) | 65 (6.5) | 102 (10.2) | 709 (71.0) | 360 (35.9) | 187 (18.6) | 307 (30.6) | 106 (10.6) | 43 (4.3) |
| Social distancing | 736 (73.3) | 215 (21.4) | 35 (3.5) | 12 (1.2) | 6 (0.6) | 856 (85.9) | 123 (12.3) | 12 (1.2) | 3 (0.3) | 3 (0.3) |
| Avoiding travel | 767 (76.6) | 171 (17.1) | 44 (4.4) | 7 (0.7) | 12 (1.2) | 835 (83.1) | 147 (14.6) | 19 (1.9) | 1 (0.1) | 3 (0.3) |
| Shelter in place/ quarantine | 582 (58.0) | 318 (31.7) | 64 (6.4) | 22 (2.2) | 18 (1.8) | 846 (84.4) | 135 (13.5) | 18 (1.8) | 0 (0) | 4 (0.4) |

All performance-belief pairs were associated significantly ( $p < 0.001$ ).

On univariate logistic regression analyses, NPI adherence was associated with a belief that NPIs would reduce personal risk of developing COVID-19 (OR 3.06, 95% CI [1.25, 7.48], p=0.014) and with a belief that the NPIs were *not* difficult to perform (OR 1.79, 95% CI [1.38, 2.31], p<0.0001). Adherence was also associated with self-described religiosity (OR 1.85, 95% CI [1.42, 2.39], p<0.0001); full-time employment (OR 1.35, 95% CI [1.02, 1.78], p=0.035); worry regarding a family member contracting COVID-19 (OR 1.47, 95% CI [1.11, 1.93], p=0.007); and belief that the media was *not* exaggerating the severity of the pandemic (OR 1.44, 95% CI [1.09, 1.91], p=0.012).

## Discussion

Most respondents stated that they are performing key NPIs, such as hand washing and social distancing, on a consistent basis, and the majority of respondents agreed that NPIs are effective in slowing the spread of COVID-19. Mask wearing and eye protection adherence and perceived efficacy lag behind other NPIs; this may be due to messaging, since at the time the survey was performed no recommendations were in place to encourage mask or face protection by the general public in the US. While some have questioned the effectiveness of school closures,^12^ it is important to maintain consistent messaging for the general public, particularly since the scientific consensus is that NPIs are effective overall.^2,5,6^ This is particularly important since beyond belief in efficacy, emotional appeals may be important in encouraging appropriate behaviors.^13^ Not surprisingly, those who believe that NPI use is not at all inconvenient are more likely to engage in NPI use, as are those that believe in the efficacy of NPIs in reducing personal risk of COVID-19 infection. Our single study incudes approximately the same number of subjects as all 16 studies included in a recent systematic review of influenza pandemic beliefs.^14^

Limitations of this survey-based study include generalizability, mitigated in part by the stratified sampling and large survey panel design; response and social desirability biases, the latter reduced by the anonymous nature of the survey; and the inability to draw causal inferences from a cross-sectional investigation.

These data highlight potential targets for public health efforts: respondents were compliant with straightforward, familiar, and heavily-encouraged NPI recommendations such as hand-washing; more onerous approaches, such as avoiding face touching, disinfecting surfaces, and wearing masks or goggles, were performed less frequently. These findings are consistent with previous research on NPIs for pandemic influenza.^6^ Changes in CDC recommendations for mask/ face coverings may impact these behaviors in the future.

An improved understanding of the drivers of refusal to engage in NPIs may help tailor messaging and increase the chances of eliciting behavioral change. NPI non-adherence is associated with both outcome expectations (NPIs are *effective*) and process expectations (NPIs are *inconvenient*). These findings have important implications for designing public health outreach efforts, where the feasibility, as well as the effectiveness, of NPIs should be stressed.

## Data Availability

The data from this study are available from the corresponding author.

